# An international comparison of age and sex dependency of COVID-19 Deaths in 2020 - a descriptive analysis

**DOI:** 10.1101/2021.03.11.21253420

**Authors:** Peter Bauer, Jonas Brugger, Franz König, Martin Posch

## Abstract

COVID-19 mortality, the number of reported COVID-19 deaths per 100,000 persons observed so far, is described in 15 European countries and the USA depending on age groups and sex for the full year 2020. It is contrasted to the corresponding historic all-cause mortality per year depending on age and sex observed in these countries. Although there are substantial differences in the age and sex dependency of COVID-19 mortality between countries, there are some common features: Exponential increase with age is a good model to describe and analyse both COVID-19 and all-cause mortality above an age of 40 years, where almost all COVID-19 death occur. Age dependency is stronger for COVID-19 mortality than for all-cause mortality, males have an excess risk compared to women which flattens off with increasing age. Also with regard to calendar time, there were differences in the age and sex dependency between countries with the common tendency that male excess risk of COVID-19 mortality was smaller in the second half of the year.

## Introduction

By December 27, 2020 more than 1.7 million COVID-19 deaths have been recorded world-wide [1]. Already early on in the pandemics it was observed that age and sex are major risk factors for COVID-19 mortality [2,17], with a higher risk for males and an exponentially increasing risk with age.

To investigate the risk patterns of COVID-19 mortality across countries, one can consider case fatality rates (CFR), defined as the number of deceased over the number of detected infections. This number, however, does not account for the dark figure of undetected infections and depends on the amount of testing and the testing strategy which substantially varies across time and between countries. In contrast, estimation of the infection fatality rate (IFR), defined as the ratio of deceased versus infected, relies on estimates of the actual number of infections, including the non-detected. While for several countries estimates based on screening tests have been provided, IFR estimates are not consistently available by ageand sex group, and are subject to limited sample sizes and limitations of the sampling procedures.

Therefore, in this paper we focus on the analysis of the population mortality rate, defined as the ratio of deceased in a certain age and sex group and the population size in that group in a certain time period. The latter quantity can be more reliably estimated based on COVID-19 deaths figures reported in different countries. However, COVID-19 population mortality rates are an overall risk measure, given by the product of the infection rate and the IFR in the respective subgroup.

While information on the number of COVID-19 death is available for many countries, definitions and data sources are not standardized. Definitions on what constitutes a COVID-19 death range from any death within 28 days after a laboratory confirmed COVID-19 infection, to deaths for which a COVID-19 infection has been identified as the cause of death in the death certificate. Analyses of excess mortality in several countries indicate that there is also a dark figure in COVID-19 deaths, although some of the excess mortality may also be due to indirect effects of the pandemic, as, for example, overburdened health systems. Furthermore, some countries report only hospital deaths which may not be representative of all COVID-19 deaths.

In this work we aim for a descriptive analysis of the age and sex specific COVID-19 population mortality rates in 15 European countries plus the US for which sufficiently detailed numbers are available over the whole year 2020 from public sources. Especially, we investigate the age gradient and impact of sex on the mortality and relate it to the corresponding patterns of all-cause mortality before the pandemic. Furthermore, we assess time trends, comparing the mortality patterns in the first and second half-year.

## Methods

Publicly available data of 14 countries on COVID-19 deaths by age and sex were taken from the National Institute for Demographic Studies (INED) [4]. In addition, Spanish COVID-19 mortality data was acquired from the webpage of the Red Nacional de Vigilancia Epidemiológica (RENAVE) [11] and Austrian COVID-19 mortality data were requested from the Austrian National Public Health Institute (Gesundheit Österreich GmbH, GÖG), which grants access to COVID data for scientific research. Furthermore, life table data of the most recent 5-year interval was used from the Human Mortality Database [3]. Data on the age- and sex-specific population size was taken either directly from the INED-dataset or from the latest information in the Human Mortality Database in case of Austria and Spain. Definitions of COVID-19 deaths vary among the countries, nevertheless we refer to each death in the dataset as COVID-19 death, regardless of the definition. The exact definition of COVID-19 death of each individual country can be obtained from the INED’s Metadata sheet [4]. See Table 1S in the supplementary material for an overview of the datasets used.

Negative binomial regression models were fitted to the proportions of registered COVID-19 deaths, both to the proportion of Covid-19 deaths in 2020 and to the proportion of all-cause deaths from the most recent life tables per country and year before 2020. Population fatality rates were analysed, defined as the number of COVID-deaths divided by the total number of persons in the respective age groups. Population sizes were taken from the most recent life tables available from the Human Mortality Database [3] in the countries. Separate analyses were performed per country. In *Model 1 COVID* the total numbers of Covid-19 deaths in 2020 were analysed in a model with the metric variable age (centred at the age of 65 and scaled in decades), the variable male sex (with female as the reference category), and the interaction age*sex with population size as an offset in each age by sex group. The same model was used to analyse all-cause mortality in the individual countries (*Model 1 ALL*). Although for all-cause mortality the age at death is available in years, the same age groups per country as in Model 1 COVID were used to avoid systematic differences arising from differences in the definition of the influence variables. Since all-cause mortality increases approximately exponentially with age from an age of 40 years on (e.g., [5,6]), the models Model 1 All, Model 1 COVID and Model 2 COVID (see below) were fitted only in the subsamples of 40 year olds and above, i.e., including only age groups which do not contain individuals below 40. Overall, this covered 98.3% of the registered COVID-19 deaths for which information on age and sex was available over all countries. To check whether this age limit has an impact on the results, in a sensitivity analysis calculations were repeated with an age limit of 30 years or above, overall covering 99.1% of COVID-19 deaths. Since age was only available in age groups (with the exception of Austria, where subject level data with age in years was available, see Table 1S in the supplement) it was approximated by the mean age of the population in the respective age group, accounting for the age distribution. Due to the exponential increase of the proportions of deaths with age, this gives slightly positively biased estimates of the number of deaths at the mean age. In a sensitivity analysis, for Austria results using ten-year age groups were compared with the results using the one-year age groups. For countries where reported age groups varied over time, the age categorization applicable to all weeks was used.

For the comparison of the age and sex dependence of COVID-19 mortality between the first and second half of the year, a second model was fitted (*Model 2 COVID)*. To this end, a data set with weekly deaths was derived from the cumulative mortality figures, calculating the increments at the reported time periods. Due to subsequent corrections, the periodically published cumulative death figures per age and sex group in some countries are not monotonic in time for some reporting periods (see Figure 1S). These numbers were monotonized by setting negative increments to 0 and reducing the death counts in the immediately preceding time period(s) by the respective amount (such that the total cumulative number of deaths per age and sex group remained unchanged). While this procedure cannot fully correct for the over-reporting (as we have no information in which time periods the over reporting actually occurred) it is expected to reduce potential bias. Weeks in which no deaths occurred were excluded from the data set. Furthermore, the factor Period was defined, with Period 1 indicating the time 1/1/2020-31/06/2020 and Period 2 from 1/7/2020 to 31/12/2020, where the first period roughly corresponds to the first wave of the pandemics. *Model 2 COVID* included the independent factors calendar week (as categorical variable, to account for the dynamics of total deaths over time), age, sex, and the two-way interactions age*sex, age*period, sex*period, and the three-way interaction age*sex*period was fitted. Period (with period 1 as the reference) was not included as a main factor because it is completely confounded with calendar week. All analysis were performed with R [18].

## Results

The cumulative total numbers of COVID-19 deaths for the 16 countries over the year 2020 can be seen in Figure 1S (supplement), where also the cumulative numbers of male and female deaths in the subgroups of age under and over 65 years (or under and over 60 years, depending on the reported age categories) of age are presented. Non-monotonicity of the cumulative numbers in individual age classes for women and men occurred in 8 of the 16 countries and were removed as described in the methods section to derive the number of deaths per week. The analysis based on *Model 1* COVID on the total numbers of COVID deaths in 2020 would not be influenced by this procedure. For the comparison of the two periods (before/after 1^st^ of July) in *Model 2 COVID* the factor week is adjusted for in the analysis by a categorical variable and it is expected that the estimates of age and sex dependence will not be substantially biased by this procedure.

Figure 1 shows the estimates of the proportion of COVID-19 deaths for females at 65 years (the estimated intercepts in the model), and the estimates of the risk ratios for age (per 10 years), sex and the twofold interaction age*sex for COVID-19 mortality of persons over 40 years of age in the year 2020 in the 16 countries. The corresponding risk ratios for the yearly all-cause mortality before 2020 estimated from the same model are shown in Figure 1 for comparison. In the supplement the numerical values per country are presented in (Table 2Sa and Table 2Sb) and the individual estimates of the risk ratios for the age dependency in females and males are shown in Figure 2S. Data and model fit are presented in Figure 2 for both types of mortality.

**Figure 1:**
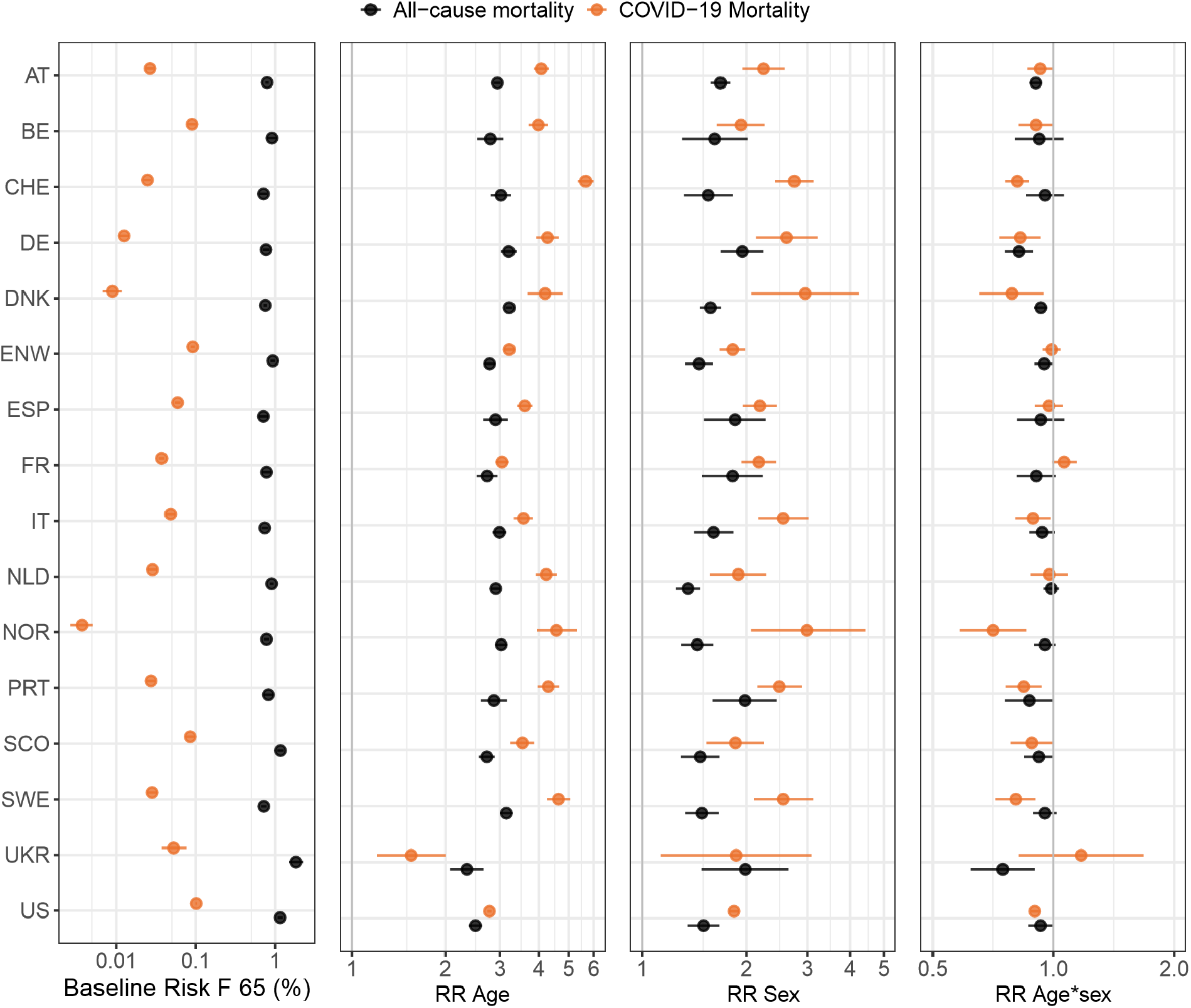
Estimates and 95% confidence interval from the negative binomial model for COVID-19 deaths in 2020 (*Model 1 COVID*) and yearly all-cause mortality before 2020 (*Model 1 ALL*) for 16 countries. Given are baseline risks (women at the age of 65), the risk ratios (RR) for age (per 10 years), for male sex and for the interaction age*sex. Results for COVID-19 mortality in orange, for all-cause mortality in black.

**Figure 2:**
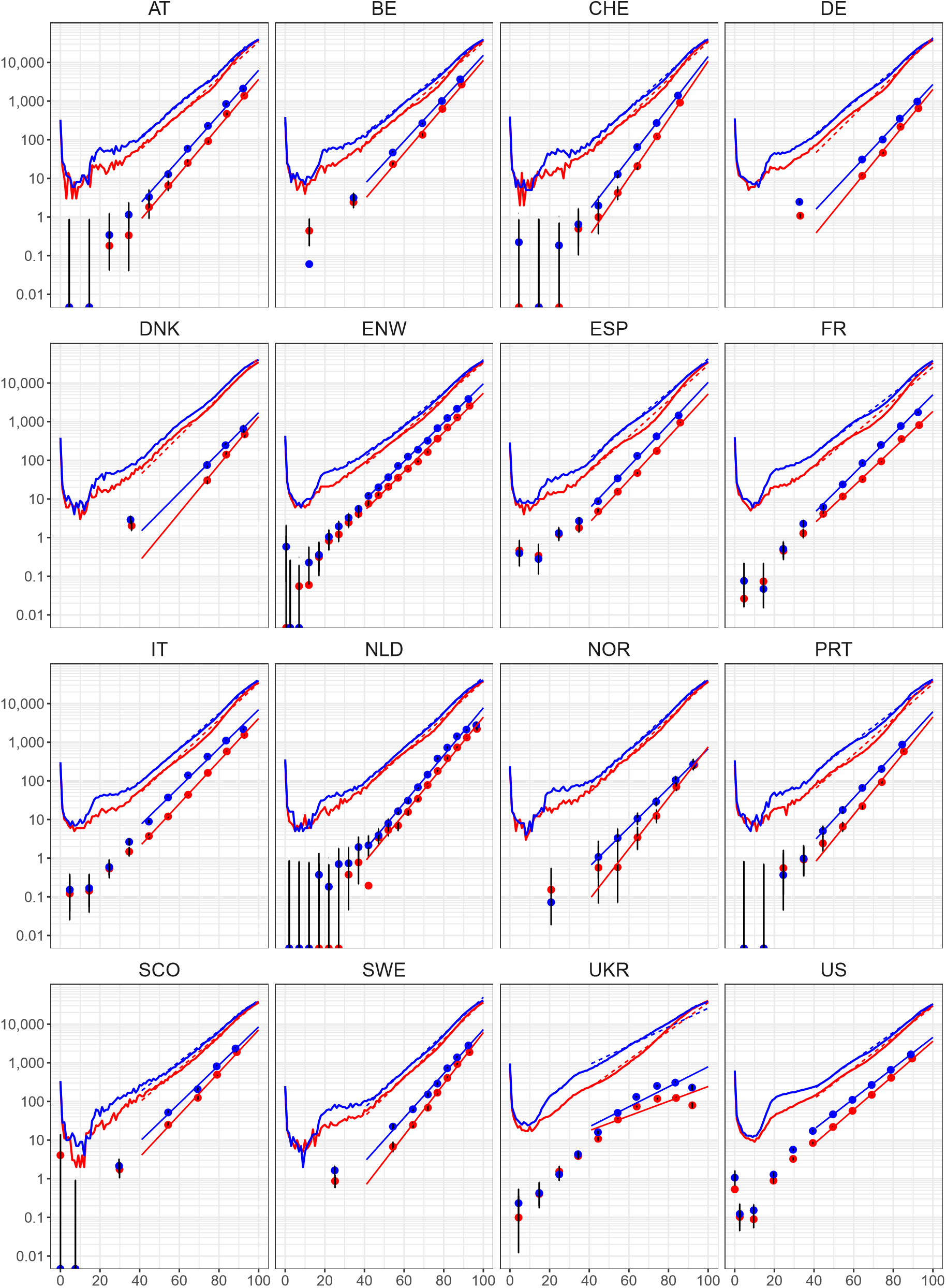
Model fit of the analysis of age and sex dependence of COVID-19 mortality and all-cause mortality. The curves on a log scales show the number of persons who normally die per 100.000 within the next year depending on age and sex, taken from all-cause mortality life tables per country before 2020, red curves for women and blue curves for men. The dots are the number of COVID-19 deaths per 100,000 plotted at the means of age in the respective age categories. The vertical lines represent 95% confidence intervals, with lower bounds cut at 0.001. The fitted model estimates for COVID-19 mortality in ages of over 40 years are shown by the solid, for all-cause mortality by the dashed lines from 40 years of age upwards.

The smallest COVID-19 mortality risk in the group of females with 65 years of age chosen as a baseline was estimated for Norway 0.004% (0.003-0.005) followed by Denmark 0.009% (0.007-0.012) and Germany 0.013% (0.011-0.015). The largest in the USA with 0.102% (0.099-0.104), with large values also in England and Wales 0.092% (0.086-0,098), Belgium 0.090% 0.080-0.101) and Scotland 0.085% (0.074-0,098). For the variable age the smallest risk ratio for COVID-19 mortality has been observed in Ukraine with, 1.55, 95% confidence interval (1.20-2.00), see (Figure 1 and Table 2Sa). This means that risk of COVID-death in women (the reference category for the variable sex) on average is 1.55 times higher if age increases by 10 years. If exponential growth applies, for females the risk to die at an age of 70 is 1.55 times higher than dying at an age of 60, and the risk of dying at an age of 80 is 1.55 times higher than to die at an age of 70. The next smallest age dependency was found in the USA with a risk ratio of 2.77 (2.73-2.82). There is a remarkable wide range of the estimates of age dependency, the largest risk ratio was found in Switzerland with 5.66 (5.36-6.00), Belgium with 3.98 (3.70-4.28) and Austria with 4.07 (3.85-4.31) were closest to the median over the 16 countries. The estimated risk of all-cause deaths for women at an age of 65 years ranged from 0.71% (0.62-0.82) in Spain and 0.72% (0.66-0.78) in Sweden up to a maximum of 1.83 (1.50-2.27) in the Ukraine followed next by Scotland with 1.17% (1.07-1.28), see Table 2Sb. With regard to the age dependency of all-cause mortality the estimates of the risk ratios per ten years vary between 2.34 (2.07 - 2.66) again lowest for Ukraine and 3.21 (3.13-3.30) highest for Denmark. For Spain with 2.90 (2.65-3.19) and the Netherlands with 2.91 (2.82-3.00) almost the same risk ratio around the median among the countries has been found. For 15 out of the 16 countries the confidence intervals for the risk ratios over a decade of age are completely disjoint between COVI-19 mortality and all-cause mortality (see Figure 1). While, not surprisingly, the age dependency of all-cause mortality before the pandemics is more homogenous over countries than the COVID-19 mortality, the numbers show that there are considerable differences in the age dependence also for all-cause mortality between the countries.

COVID-19 mortality for males (at the age of 65) was consistently larger for males as compared to females. It was closest to females in England and Wales with a risk ratio of only 1.82 (1.68-1.98) for males versus females, but with 2.99 (2.06-4.42) the difference was largest in Norway (although with a wide confidence interval). Again this is a remarkable spread of estimates over countries. The estimated risk ratios 2.19 (1.95-2.45) for Spain and 2.24 (1.95-2.59) for Austria were closest to the median over the 16 countries.

Also for all-cause mortality, the risk ratios between men and women at the age of 65 are consistently above one but they are consistently smaller than for COVID-19 mortality except for Ukraine, with risk ratios ranging from 1.35 (1.25-1.47) in the Netherlands to 1.98 (1.48-2.65) in the Ukraine, but also Portugal and Germany being close to this high estimate (Fig.1 and Table 3s). For all-cause mortality the risk ratios of 1.58 (1.47-1.69) in DNK and 1.61 (1.41-1.83) in Italy were closest to the median over the countries. When considering the interaction between age and sex there is a tendency that male and female mortality rates come closer to each other as over the age increases, for COVID-19 mortality the risk ratio of male versus females over a decade of age reduces most pronounced (by a factor of 0.71 (0.58-0.86)) in Norway. In 10 of the 16 countries the 95%-confidence intervals for the risk ratio connected with corresponding to the interaction age *sex are completely below one, all the 6 other confidence intervals cover the value of one (Figure 1). For all-cause mortality, all estimated risk ratios for the interaction between age and sex are below one, indicating the flattening of the excess risk of males with age. Comparing the interaction effects between all-cause mortality with COVID-19 mortality, there is no clear trend, in 9 of the countries the corresponding estimates being larger for all-cause mortality, in 7 countries they are smaller than for COVID-19 mortality. In Figure 2S the estimated age dependency in terms of the risk ratio per decade of age for COVID-19 mortality and all-cause mortality are contrasted between females and males. By the way, it is somewhat surprising that only in 5 countries the corresponding confidence intervals for the interaction term in all-cause mortality completely fall below one (Tab.2Sb). This may change if younger age groups are included into the calculations where sex differences in all-cause mortality are known to be larger. Indeed, in a sensitivity analysis when taking 30 years as a lower limit of age the estimates for the interaction age*sex slightly shift towards smaller values, now in already 9 countries the corresponding confidence intervals are located completely below 1. Other effect estimates for COVID-19 and all-cause mortality form the sensitivity analysis are not noticeably changing the trends seen in the primary analyses with an age limit of 40. Only in the Ukraine, the age effect for COVID-19 mortality turned out to be increased for the sample with the lower age limit of 30 since here exponential age dependency is flattening off for higher ages. The deviating features of the data registered in the Ukraine also result in a poorer fit of the model and wider confidence intervals for some of the effect estimates (see Figure 2). In a further sensitivity analysis with Austrian data, we looked how much results change, if the models were be fitted with one year age intervals (available in Austria) as compared to 10 years as available for many other countries. No relevant deviations between the estimates arising from the differences in length of the age categories were found (see Table2Sa).

Overall, the fit of the models appears to be good, the fit for all-cause mortality above 40 years being hardly distinguishable from the life table data (Figure 1). Hence exponential growth of the risk of dying with increasing age is ascertained to be a good model for describing and analysing both COVID-19 mortality and all-cause mortality over 40 years of age. There are, however, considerable differences in the characteristic of the age and sex dependency of COVID-19 mortality estimates between countries and several confidence intervals of the risk ratios for age and sex do not overlap between countries. This heterogeneity is a strong argument in favour of performing per country analyses. In Norway, the COVID-19 mortality in 2020 is substantially below the all-cause mortality per year, whereas in Belgium and England and Wales the curves are closer. In the USA with the lowest age dependency besides the Ukraine, the COVID-19 and all-cause mortality curves seem to be almost parallel. Only in the Ukraine the reported numbers show a deviating behaviour and COVID-19 mortality appears to be not monotonically increasing with age, leading to a poor model fit. For many countries, the low numbers of registered COVID-19 death at young ages make it difficult to infer any convincing results for these age groups, as may be seen from the wide confidence intervals in Figure 2. However, for large countries such as the USA there seems be a touch of similarity to the age dependency of all-cause mortality even in these low ages.

Figure 3 shows a scatterplot of the estimates of the risk ratios for the age dependency in males and females of COVID-19 mortality per versus the corresponding estimates for all-cause mortality per country and year, computed from *Model 1 COVID and Model 1 ALL*. Overall, despite the existing heterogeneity of results there is a considerable correlation between the two risk ratios estimated (Spearman Rho for males and females together r=0.72, n=32). When looking separately at males (Spearman Rho 0.50, n=16) and females (Spearman Rho r=0.79, n=16) this may be taken as a minor hint on a possibly less stringent correlation of the estimates for males.

**Figure 3:**
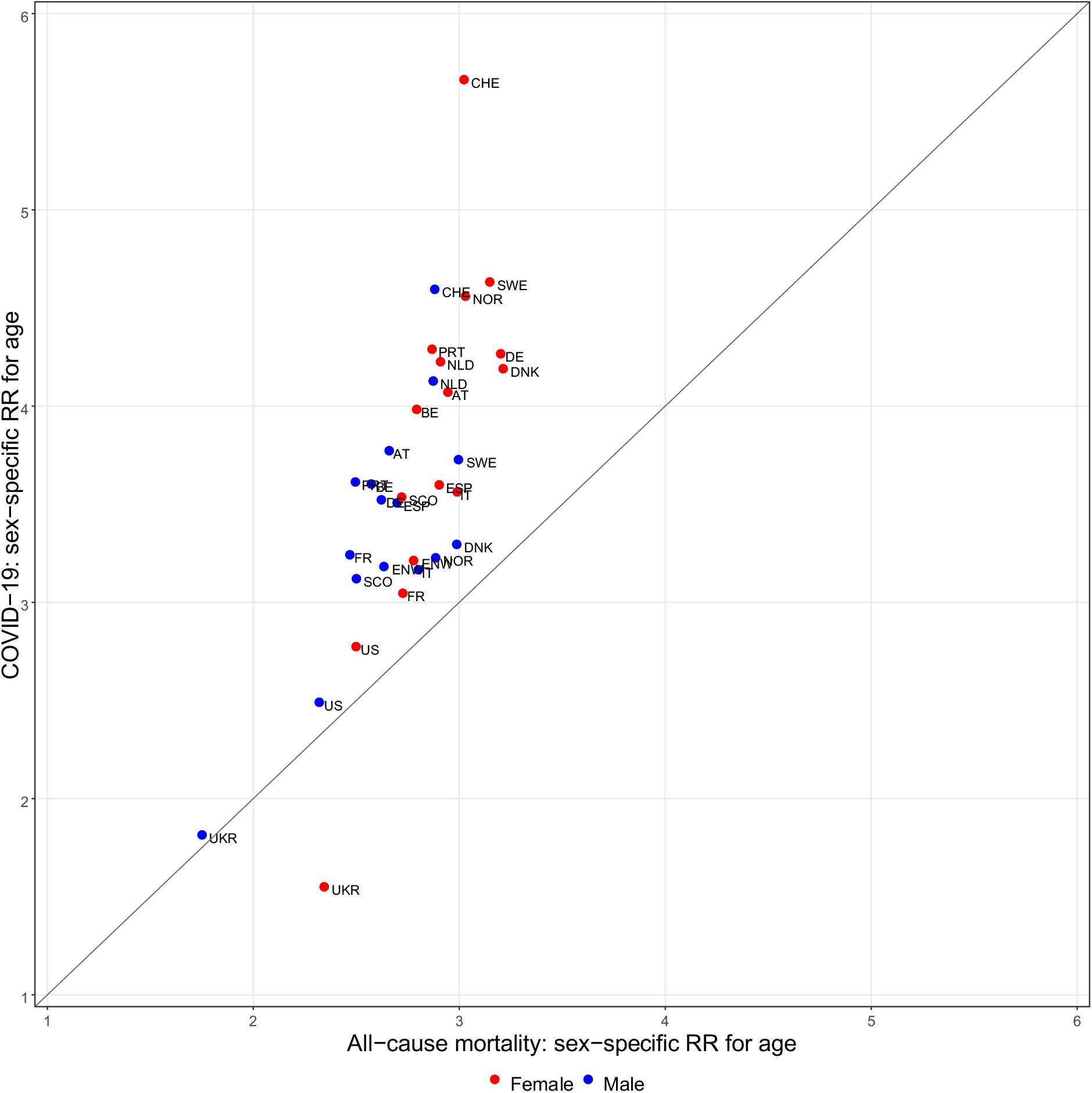
Scatterplot of the risk ratios for age (per 10 years of age) of males and females estimated for COVID-19 mortality in 2020 versus the corresponding risk ratios estimated for all-cause mortality before 2020. The risk ratios are estimated from *Model 1 COVID* and *Model 1 All*, Red dots: females, blue dots: males.

At a next level of analysis, in *Model 2 COVID*, in addition to the variables already used in *Model 1*, the factor calendar week was included and interactions of the factor period (second versus first half of the year) with age and sex up to the threefold interaction. The main factor for period cannot be estimated in a model including the factor calendar weeks. However, *Model 2* allows to investigate whether there have been changes in age and sex dependencies of COVID-19 mortality during 2020. The estimates of the main effects age and sex and of the interactions of age*period, sex*period age*sex*period are shown in forest plots per country in Figure 4. There were differences of the age dependencies of COVID-19 mortality between the two half years among countries. In Belgium, Scotland and the USA the age dependency was less accentuated in the second half year with an estimate of the risk ratio for the interaction of age*period below 1 and also the confidence interval for the effect fully falling below 1. In Switzerland, Denmark and France the corresponding estimates of the risk ratio were above 1 and with confidence intervals fully falling above 1, see Figure 4. This is an extremely unlikely pattern of estimates if in all countries no differences for age dependency between periods would exist at all.

**Figure 4:**
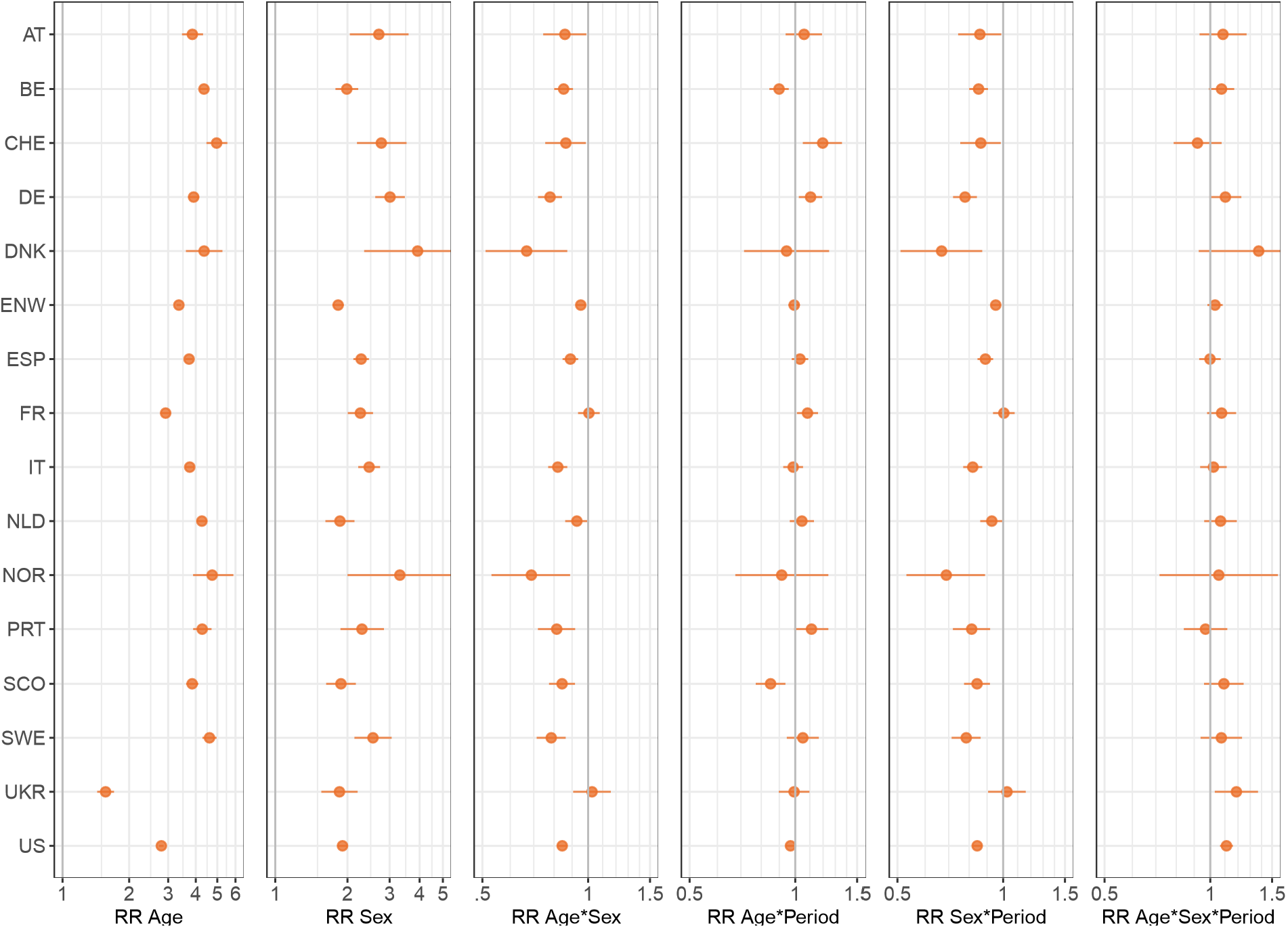
Estimates and 95% confidence intervals for the risk ratios of age, sex, period, the twofold interactions age*sex, age*period, sex*period and the threefold interaction age*sex*period for COVID-19 mortality from *Model* 2 COVID.

The only convincingly stable result over countries was that in the second half of the year 2020 COVID-19 mortality was closer between men and women than in the first half of the year. In thirteen countries the confidence intervals of the corresponding risk ratios for the interaction period*sex are completely below 1, see Figure 4, meaning that the difference between 65 year old men and women decreased in the second period. When looking at the threefold interaction age*sex*period the estimates in 14 of the 16 are above 1, however, only two of them with a confidence interval fully above 1. This may be taken as a hint that the difference in age dependency between males and females was attenuated in the second period of 2020.

## Discussion

The main purpose of this analysis was to describe the age and sex dependencies of COVID-19 deaths as documented in 15 European countries and the USA. Because reliable numbers on the distribution of infected persons per age and sex are not available across countries, estimation of age and sex specific infection fatality rates per country is not feasible without strong assumptions. Therefore we looked at population COVID-19 fatality rates based on the age and sex distributions of the population which are available from official sources in these countries. If only a small proportion of the population is infected, these population fatality rates are substantially lower than the infection fatality rates. This may cause confusion in the public perception if not communicated appropriately. Population COVID-19 fatality rates combine infection rates and infection fatality rates. This implies that age and sex dependencies found for the population fatality rate cannot be directly extrapolated to corresponding dependencies for the infection fatality rates unless a constant infection rate over age groups and sex per country applies.

An important result of the analysis is that generally the percentage of increase in COVID-19 mortality per 10 years of increasing age was larger than the percentage of increase of all-cause mortality over 10 years of age in the countries before the pandemics. Roughly, in terms of the median over countries, the risk of COVID death in 2020 for an increase in age of ten years increased by a factor of around 4, whereas for all-cause mortality it only increases by a factor of slightly below 3. Also the excess of COVID-19 mortality in men as compared to women in terms of risk ratios was more accentuated for COVID-19 mortality than for all-cause mortality, here the median risk ratio for COVID-19 death in males as compared to females at an age of 65 years was about 2.2 as compared to 1.6 for all-cause mortality. In the majority of the countries, the age dependency of the excess of male risk in COVID-19 mortality decreased with age, men coming closer to women in older age groups. This trend is known also for all-cause mortality and we found no indication that the magnitude of the trend would differ much between COVID-19 and all-cause mortality over countries. Whereas the absolute numbers of COVID-19 death for males and females are different for lower age groups, they are almost identical in higher age groups(see Figure 1S). So at the first glance one might expect larger interaction effects between age and sex. Note however, that in higher age groups the numbers at risk are larger for females than for males. From early data in China, South Korea, Italy, Spain, France, Germany, England and Wales, and the United States up to April 2020 without differentiation for sex it was found that all-cause mortality is increasing exponentially at a constant rate of about 10% per year of age and COVID-19 mortality increases at about 11% per year of age [9], which is not too far away from our estimates for yearly increases of around 12% and 15% corresponding to ten years risk ratios of 3 and 4. In an analysis of data from eight European countries up to May 2020 differentiating for sex the progression ratios per ten years of age of COVID-19 mortality has been estimated as 3.3 and 3.4 for females and males respectively [6] which does not indicate any narrowing of the excess risk of men with increasing age. Ahrenfeldt et al. [7] for 10 European countries reported crude relative risks for males in four age groups from data collected latest up to June 29^th^, 2020. They found the smallest differences between male and females in the age group over 85 years of age.

In an analysis covering data of two waves of the pandemics in 2020 in 14 countries the age distribution of COVID-19 deaths of males and females together has been fairly similar in the second versus the first wave [8]. We found differences in the age dependency of COVID-19 mortality between the two half year periods in 2020 among the 16 countries. For some countries the age dependency being more accentuated in the second half of the year for others more in the first half. The only consistent period effect of COVID-19 mortality over the calendar year 2020 in our analysis was the decrease of excess mortality of males in the second half of the year. To explain this effect, one would have to know whether there have been some common systematic differences between females and males over countries in contacts, behaviour and infections between females and males over the pandemics, and if or why such differences may have levelled out during the pandemic? It is not surprising, that estimates of the risk ratios for age quite well correlate between COVID-19 mortality and all-cause mortality over countries: Older persons have a much larger risk to die in years outside the pandemics, so they may also have a much higher risk to die from or with the infection during a pandemic. Such a correlation has been reported also in [9] where, however, sex was not considered as factor.

While consistent in the qualitative trends, the age and sex dependency of COVID-19 mortality varied quite noticeably between countries. We did not try to speculate about reasons for such differences since there are so many possible sources of variability as, e.g., differences in the measures against the pandemics, in the willingness of the populations to adhere to advices, in testing, in the health system, in the health awareness of the populations, in elderly care, in the registration procedures of COVID-19 deaths, in the political situation, and so on. Interpretation should be tried locally for the respective countries utilizing detailed information on specific events occurring and measures taken during the pandemics within each country, but also considering geographic location [10]. For the interpretation one has to keep in mind that there are also differences in all-cause mortality between countries. However, demographic characteristics between countries have been successfully used to explain differences in mortality between countries [12,13,16]. Furthermore, also other demographic factors as lower income or lower education have been identified as risks [14,15]. We believe that the way we looked at the data allows for a straightforward comparison of the age and sex dependency of COVID-19 mortality with the age and sex dependency of all-cause mortality. In addition, the identification of time trends in the age and sex distribution of COVID-19 mortality can provide important information for the monitoring of measures to fight the pandemics.

## Data Availability

All data (with the exception of the data for Austria) are publicly available. The data for Austria has been obtained from the GOEG Data Platform COVID-19 https://datenplattform-covid.goeg.at/ For details see the article.

## Author Contributions

All authors designed the study and wrote the manuscript. JB performed the data analysis and the figures. All authors reviewed and revised the manuscript.

## Additional Information

The authors declare that there is no conflict of interest.

