## Supplementary Material for "An international comparison of age and sex dependency of COVID-19 Deaths in 2020 - a descriptive analysis"

**Peter Bauer, Jonas Brugger, Franz König, Martin Posch**

Section for Medical Statistics, Center for Medical Statistics, Informatics and Intelligent Systems, Medical University of Vienna, Spitalgasse 23, 1090 Vienna, Austria

| Country | Population Size | COVID-19 Deaths | % of COVID-19 deaths with missing age or sex | Age categories | Average number of reports per Week | Weeks without report | Data set |
|---|---|---|---|---|---|---|---|
| AT | 8,859,650 | 5,306 | 0.00 | 1-year intervals | 7.00 | - | Data set provided by GÖG |
| BE | 11,492,641 | 19,163 | 0.19 | 0-24, 25-44, 45-64, 65-74, 75-84, 85+ | 6.93 | - | Deaths_by_occurrence |
| CHE | 8,582,905 | 6,777 | 0.02 | 0-9, 10-19, 20-29, 30-39, 40-49, 50-59, 60-69, 70-79, 80+ | 3.18 | 32,33,36,37 | Dataset name missing (see table caption) |
| DE | 83,019,213 | 26,964 | 0.16 | 0-59, 60-69, 70-79, 80-89, 90+ | 4.62 | 38 | SSI_by age and sex_Data |
| DNK | 5,822,763 | 1,070 | 11.13 | <70, 70-79, 80-89, 90+ | 5.30 | - | Daily Report RKI_Data |
| ENW | 59,115,809 | 73,730 | 0.00 | <1, 1-4, 5-9, 10-14, 15-19, 20-24, 25-29, 30-34, 35-39, 40-44, 45-49, 50-54, 55-59, 60-64, 65-69, 70-74, 75-79, 80-84, 85-89, 90+ | 1.00 | - | ONS_WeeklyOccurrenceDeaths |
| ESP | 46,937,063 | 50,610 | 0.01 | 0-9, 10-19, 20-29, 30-39, 40-49, 50-59, 60-69, 70-79, 80+ | 7.00 | - | Data obtained from RENAVE [11] |
| FR | 67,063,703 | 42,615 | 1.06 | 0-9, 10-19, 20-29, 30-39, 40-49, 50-59, 60-69, 70-79, 80-89, 90+ | 6.78 | - | SpF_by age and sex_HospitalData |
| IT | 60,359,546 | 67,537 | < 0.01 | 0-9, 10-19, 20-29, 30-39, 40-49, 50-59, 60-69, 70-79, 80-89, 90+ | 1.36 | - | Combined_Information |
| NLD | 17,282,163 | 10,633 | 0.00 | 0-4, 5-9, 10-14, 15-19, 20-24, 25-29, 30-34, 35-39, 40-44, 45-49, 50-54, 55-59, 60-64, 65-69, 70-74, 75-79, 80-84, 85-89, 90-94, 95+ | 2.90 | - | RIVM_Data |
| NOR | 5,367,580 | 429 | 0.00 | <40, 40-49, 50-59, 60-69, 70-79, 80-89, 90+ | 4.68 | - | FHI_Deaths by age and sex |
| PRT | 10,276,617 | 6,674 | 0.05 | 0-9, 10-19, 20-29, 30-39, 40-49, 50-59, 60-69, 70-79, 80+ | 6.89 | 34 - 47 | min-sau_Data |
| SCO | 5,463,300 | 6,298 | 0.00 | 0, 1-14, 15-44, 45-64, 65-74, 75-84, 85+ | 1.00 | - | NRS Age & Sex |
| SWE | 10,327,553 | 7,802 | 0.00 | 0-49, 50-59, 60-69, 70-74, 75-79, 80-84, 85-89, 90+ | 1.12 | - | PHAS_Data |
| UKR | 41,732,779 | 16,446 | 0.00 | 0-9, 10-19, 20-29, 30-39, 40-49, 50-59, 60-69, 70-79, 80-89, 90+ | 6.60 | - | Deaths by age and sex |
| US | 327,167,434 | 291,749 | < 0.01 | 0, 1-4, 15-24, 25-34, 35-44, 45-54, 5-14, 55-64, 65-74, 75-84, 85+ | 1.00 | - | CDC_Data |

Table 1S Overall population, COVID-19 deaths in 2020, % of deaths reported for which no information age and/or sex was available, age groups for which deaths were reported, average number of reports per weeks during the reporting period, weeks in which no updated figures are available, and data source. With the exception of Austria and Spain, the data source refers to the name of the data sheet obtained from INED (see the methods section in the main manuscript). Data from Switzerland was obtained from INED, but Switzerland is not listed in their metadata table.

**a) COVID-19 mortality in 2020**

| Country | Baseline Risk F 65 (%) | RR Age | RR Sex | RR Age*Sex |
|---|---|---|---|---|
| AT | 0.027 (0.024-0.030) | 4.07 (3.85-4.31) | 2.24 (1.95-2.59) | 0.93 (0.86-1.00) |
| | *0.028 (0.024-0.032)* | *4.12 (3.83-4.45)* | *2.18 (1.83-2.61)* | *0.94 (0.85-1.04)* |
| BE | 0.090 (0.080-0.101) | 3.98 (3.70-4.28) | 1.93 (1.64-2.26) | 0.90 (0.82-1.00) |
| CHE | 0.025 (0.022-0.028) | 5.66 (5.36-6.00) | 2.75 (2.43-3.13) | 0.81 (0.76-0.87) |
| DE | 0.013 (0.011-0.015) | 4.27 (3.92-4.64) | 2.62 (2.13-3.22) | 0.83 (0.73-0.93) |
| DNK | 0.009 (0.007-0.012) | 4.19 (3.67-4.78) | 2.96 (2.07-4.25) | 0.79 (0.65-0.95) |
| ENW | 0.092 (0.086-0.098) | 3.21 (3.10-3.34) | 1.82 (1.68-1.98) | 0.99 (0.94-1.04) |
| ESP | 0.059 (0.054-0.064) | 3.60 (3.40-3.81) | 2.19 (1.95-2.45) | 0.97 (0.90-1.06) |
| FR | 0.037 (0.034-0.041) | 3.05 (2.90-3.20) | 2.17 (1.93-2.44) | 1.06 (0.99-1.14) |
| IT | 0.048 (0.043-0.055) | 3.56 (3.32-3.83) | 2.56 (2.17-3.03) | 0.89 (0.80-0.98) |
| NLD | 0.029 (0.025-0.033) | 4.23 (3.91-4.58) | 1.89 (1.57-2.28) | 0.98 (0.88-1.09) |
| NOR | 0.004 (0.003-0.005) | 4.56 (3.94-5.32) | 2.99 (2.06-4.42) | 0.71 (0.58-0.86) |
| PRT | 0.028 (0.024-0.031) | 4.29 (3.96-4.66) | 2.49 (2.15-2.89) | 0.84 (0.76-0.93) |
| SCO | 0.085 (0.074-0.098) | 3.54 (3.24-3.86) | 1.86 (1.53-2.25) | 0.88 (0.78-1.00) |
| SWE | 0.028 (0.024-0.033) | 4.63 (4.25-5.06) | 2.56 (2.10-3.12) | 0.80 (0.72-0.90) |
| UKR | 0.052 (0.038-0.077) | 1.55 (1.20-2.00) | 1.87 (1.13-3.08) | 1.17 (0.82-1.68) |
| US | 0.102 (0.099-0.104) | 2.77 (2.73-2.82) | 1.84 (1.78-1.90) | 0.90 (0.88-0.92) |

**b) all-cause mortality**

| Country | Baseline Risk F 65 in % | RR Age | RR Sex | RR Age*Sex |
|---|---|---|---|---|
| AT | 0.789 (0.754-0.827) | 2.94 (2.87-3.02) | 1.68 (1.58-1.79) | 0.90 (0.87-0.94) |
| BE | 0.909 (0.782-1.066) | 2.79 (2.53-3.08) | 1.62 (1.30-2.02) | 0.92 (0.80-1.06) |
| CHE | 0.711 (0.635-0.800) | 3.02 (2.80-3.26) | 1.55 (1.32-1.83) | 0.95 (0.85-1.06) |
| DE | 0.764 (0.692-0.846) | 3.20 (3.02-3.39) | 1.94 (1.69-2.24) | 0.82 (0.75-0.89) |
| DNK | 0.750 (0.713-0.789) | 3.21 (3.13-3.30) | 1.58 (1.47-1.69) | 0.93 (0.90-0.97) |
| ENW | 0.931 (0.871-0.996) | 2.78 (2.67-2.89) | 1.46 (1.33-1.60) | 0.95 (0.90-1.00) |
| ESP | 0.709 (0.615-0.822) | 2.90 (2.65-3.19) | 1.85 (1.51-2.27) | 0.93 (0.81-1.07) |
| FR | 0.776 (0.675-0.899) | 2.73 (2.52-2.95) | 1.82 (1.49-2.23) | 0.91 (0.81-1.01) |
| IT | 0.736 (0.672-0.808) | 2.99 (2.84-3.15) | 1.61 (1.41-1.83) | 0.94 (0.87-1.01) |
| NLD | 0.901 (0.851-0.955) | 2.91 (2.82-3.00) | 1.35 (1.25-1.47) | 0.99 (0.94-1.03) |
| NOR | 0.776 (0.718-0.839) | 3.03 (2.90-3.17) | 1.44 (1.29-1.61) | 0.95 (0.90-1.01) |
| PRT | 0.820 (0.708-0.958) | 2.87 (2.60-3.16) | 1.98 (1.60-2.45) | 0.87 (0.75-1.00) |
| SCO | 1.168 (1.067-1.282) | 2.72 (2.56-2.89) | 1.47 (1.29-1.68) | 0.92 (0.84-1.00) |
| SWE | 0.718 (0.663-0.778) | 3.15 (3.00-3.30) | 1.49 (1.33-1.67) | 0.95 (0.89-1.02) |
| UKR | 1.832 (1.503-2.266) | 2.34 (2.07-2.66) | 1.98 (1.48-2.65) | 0.75 (0.62-0.90) |
| US | 1.155 (1.072-1.247) | 2.50 (2.38-2.63) | 1.50 (1.35-1.67) | 0.93 (0.86-1.00) |

**Table 2S:** Estimates and 95% confidence interval from the negative binomial model for COVID-19 mortality in 2020 (Model 1 COVID, Table a) and all-cause mortality (Model 1 ALL, Table b). Baseline risk (in %) for women at 65 years of age, risk ratios (RR) for age per 10 years, male sex, and the interaction age*sex in the 16 countries. The second row for Austria (in italics) shows the estimate of the model fitted with age categorized in 10 year age intervals.

| Country | RR Age | RR Sex | RR Age*Sex | RR Age*Period | RR Sex*Period | RR Age*Sex*Period |
|---|---|---|---|---|---|---|
| AT | 3.86 (3.46-4.31) | 2.70 (2.05-3.60) | 0.86 (0.74-0.99) | 1.06 (0.94-1.19) | 0.86 (0.74-0.99) | 1.09 (0.93-1.27) |
| BE | 4.35 (4.16-4.56) | 1.99 (1.78-2.22) | 0.85 (0.80-0.91) | 0.90 (0.84-0.96) | 0.85 (0.80-0.91) | 1.08 (0.99-1.17) |
| CHE | 4.97 (4.47-5.55) | 2.77 (2.19-3.53) | 0.86 (0.75-0.99) | 1.20 (1.05-1.36) | 0.86 (0.75-0.99) | 0.92 (0.79-1.08) |
| DE | 3.91 (3.69-4.14) | 3.01 (2.61-3.47) | 0.78 (0.72-0.84) | 1.11 (1.02-1.19) | 0.78 (0.72-0.84) | 1.10 (0.99-1.23) |
| DNK | 4.36 (3.61-5.28) | 3.92 (2.35-6.61) | 0.67 (0.51-0.87) | 0.94 (0.71-1.25) | 0.67 (0.51-0.87) | 1.37 (0.93-2.04) |
| ENW | 3.35 (3.27-3.44) | 1.83 (1.73-1.93) | 0.95 (0.92-0.98) | 0.99 (0.96-1.03) | 0.95 (0.92-0.98) | 1.03 (0.98-1.08) |
| ESP | 3.73 (3.59-3.88) | 2.28 (2.12-2.46) | 0.89 (0.85-0.94) | 1.03 (0.98-1.09) | 0.89 (0.85-0.94) | 1.00 (0.93-1.07) |
| FR | 2.92 (2.78-3.08) | 2.26 (2.01-2.56) | 1.00 (0.94-1.08) | 1.08 (1.01-1.16) | 1.00 (0.94-1.08) | 1.08 (0.98-1.18) |
| IT | 3.75 (3.58-3.94) | 2.46 (2.22-2.74) | 0.82 (0.77-0.87) | 0.99 (0.92-1.05) | 0.82 (0.77-0.87) | 1.02 (0.94-1.11) |
| NLD | 4.26 (4.03-4.51) | 1.86 (1.62-2.14) | 0.93 (0.86-1.00) | 1.04 (0.96-1.13) | 0.93 (0.86-1.00) | 1.07 (0.96-1.19) |
| NOR | 4.74 (3.89-5.87) | 3.31 (2.00-5.66) | 0.69 (0.53-0.89) | 0.91 (0.68-1.24) | 0.69 (0.53-0.89) | 1.06 (0.72-1.56) |
| PRT | 4.27 (3.89-4.71) | 2.30 (1.87-2.84) | 0.81 (0.72-0.92) | 1.11 (0.99-1.24) | 0.81 (0.72-0.92) | 0.97 (0.84-1.12) |
| SCO | 3.85 (3.62-4.11) | 1.88 (1.63-2.17) | 0.84 (0.77-0.92) | 0.85 (0.77-0.94) | 0.84 (0.77-0.92) | 1.09 (0.96-1.24) |
| SWE | 4.61 (4.29-4.97) | 2.55 (2.14-3.06) | 0.78 (0.71-0.86) | 1.05 (0.95-1.17) | 0.78 (0.71-0.86) | 1.08 (0.94-1.23) |
| UKR | 1.56 (1.43-1.71) | 1.85 (1.56-2.21) | 1.03 (0.91-1.16) | 0.99 (0.90-1.10) | 1.03 (0.91-1.16) | 1.19 (1.03-1.37) |
| US | 2.79 (2.73-2.86) | 1.91 (1.81-2.00) | 0.84 (0.82-0.87) | 0.97 (0.94-1.00) | 0.84 (0.82-0.87) | 1.11 (1.07-1.16) |

**Table 3S:** Estimates and 95% confidence interval from the negative binomial model for COVID-19 mortality in 2020 (Model 2 COVID) based on weekly death counts. Risk ratios (RR) for age per 10 years, male sex, and the interactions age*sex, age*period, sex*period, and age*sex*period in the 16 countries. Estimates for the factor calendar week not shown.

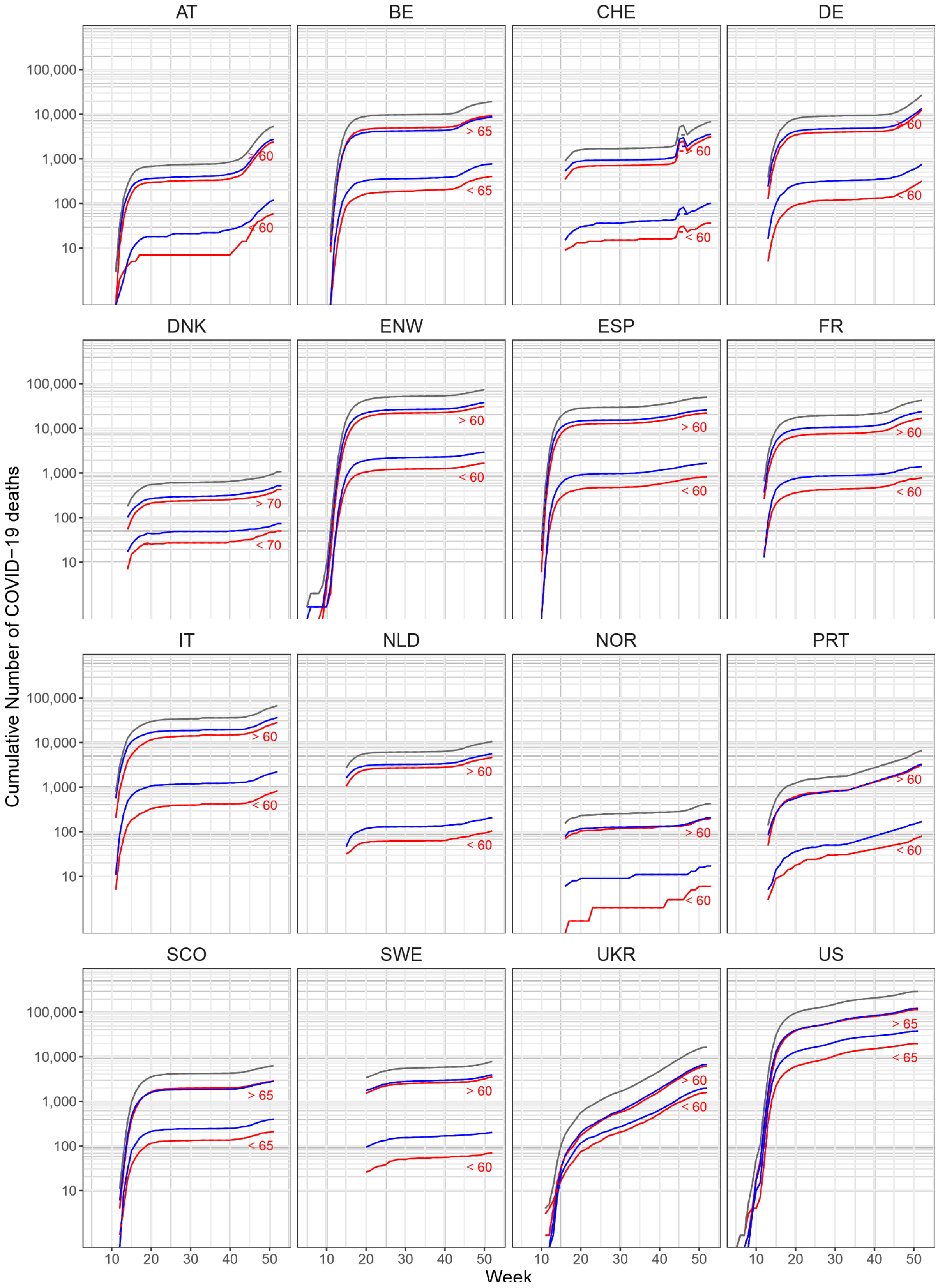


**Figure 1S:** Logarithm of the cumulative numbers of COVID-19 deaths over the calendar weeks of 2020 in 16 countries. Black: total numbers; red: female, blue: male, both for deaths <=60 (or <=65) and >60 (or >65) years of age. For some countries such as Switzerland there seem to be data errors and the monotonized by age group (see the methods section in the main manuscript) are given by dashed lines.

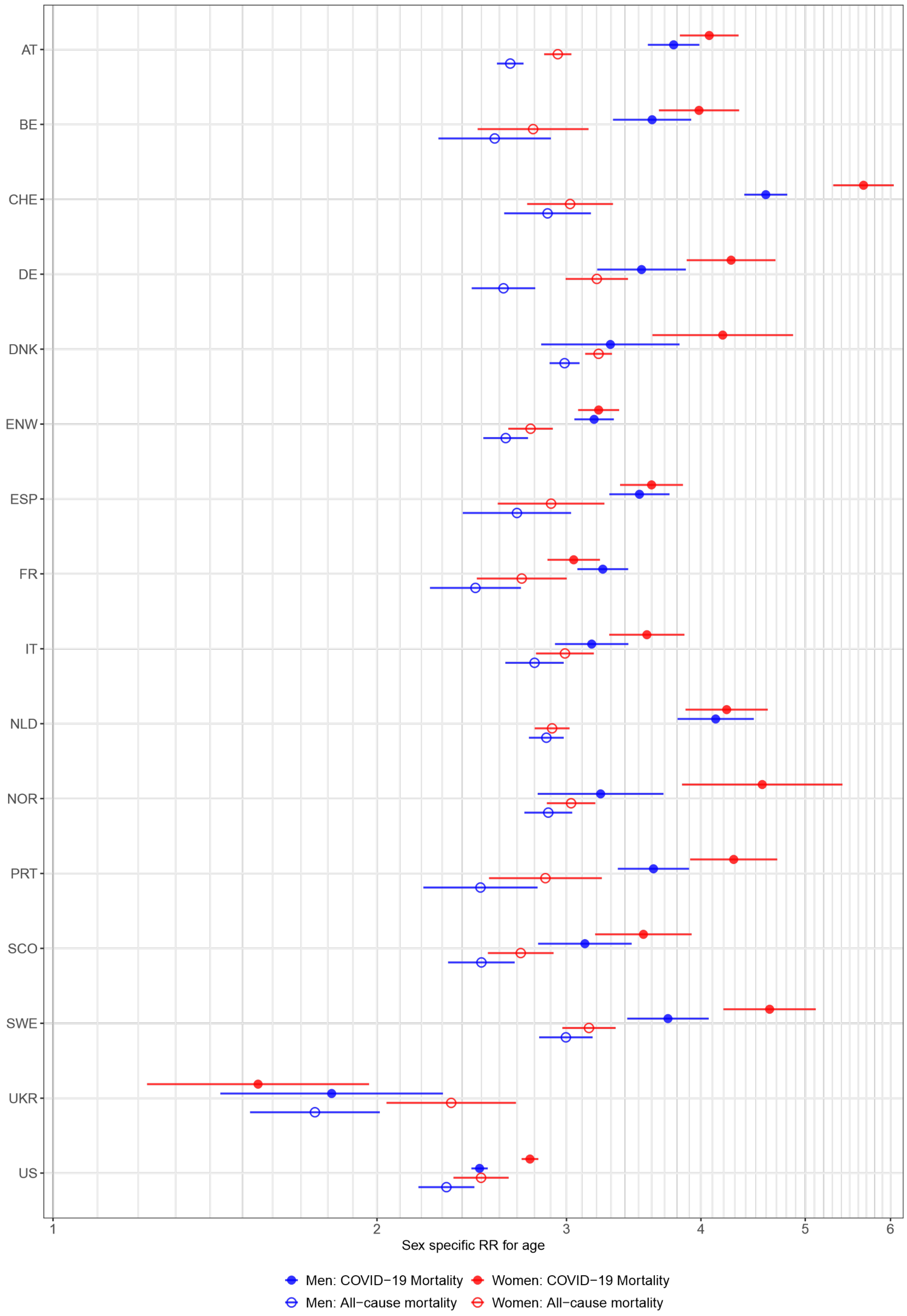


**Figure 2S:** Risk ratios and 95%-confidence intervals for the age (per 10 years) of COVID-19 mortality (full circles) and all-cause mortality (empty circles) in females (red) and males (blue) per country obtained from Model 1 COVID and Model 1 ALL.

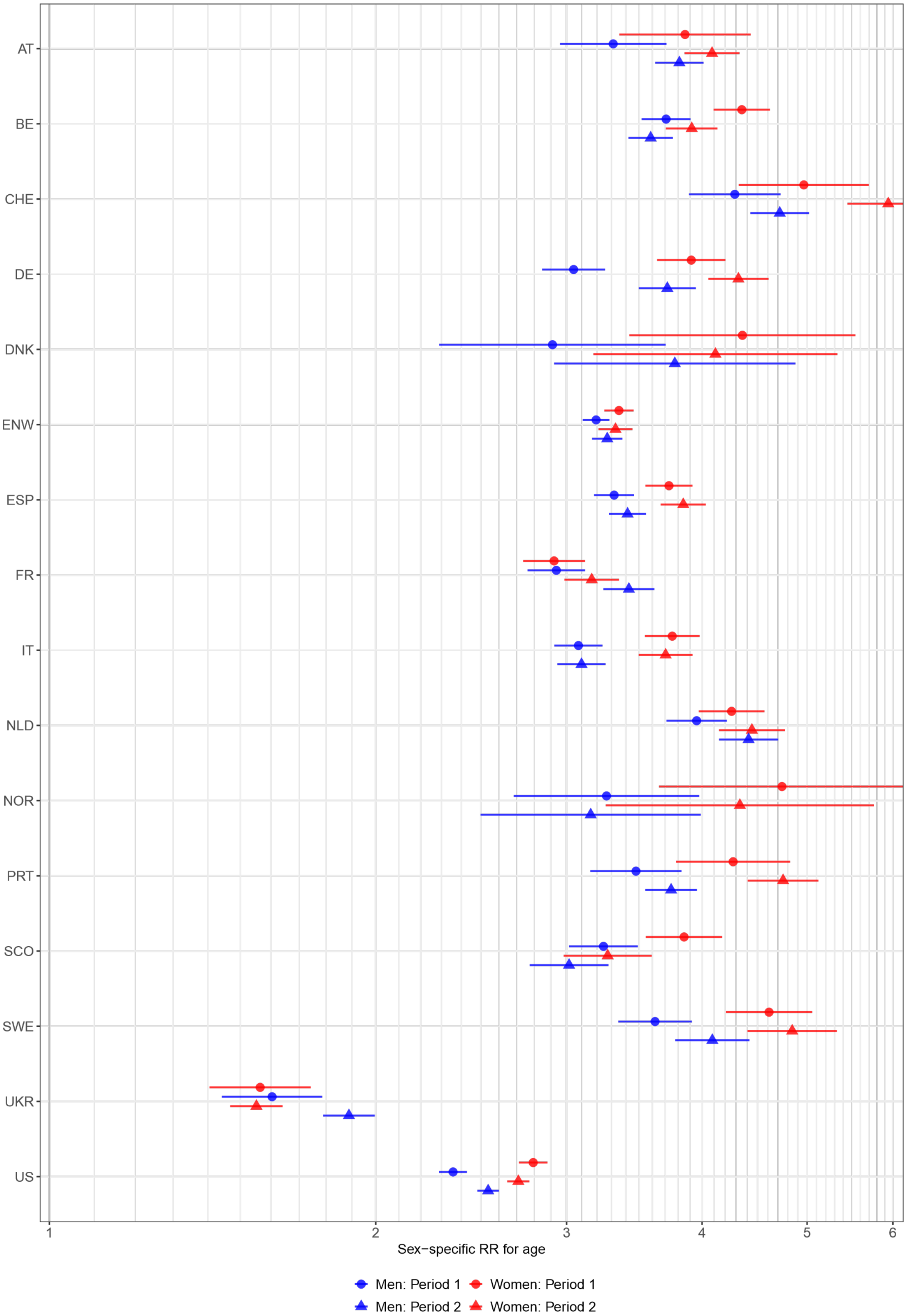


**Figure 3S:** Sex-specific Risk ratios and 95%-confidence intervals for the age (per 10 years) of COVID-19 mortality separately for the first (full circles) and second period (full triangles) in females (red) and males (blue) per country obtained from Model 2 COVID.
